# Increasing power in language genetics with Lingo: a web-based digital phenotyping platform

**DOI:** 10.1101/2024.03.29.24305034

**Authors:** Lucas G. Casten, Tanner Koomar, Muhammad Elsadany, Ying Hsu, Emily Kelly, Gabriella Snyder, Trevor Jackson, Kaleb McKone, Mahesh Sasidharan, J. Bruce Tomblin, Jacob J. Michaelson

**Affiliations:** Department of Psychiatry, University of Iowa, Iowa City, IA; Hawkeye Intellectual and Developmental Disabilities Research Center (Hawk-IDDRC), University of Iowa, Iowa City, IA; Department of Psychological and Brain Sciences, University of Iowa, Iowa City, IA; Department of Communication Sciences and Disorders, University of Iowa, Iowa City, IA; Iowa Neuroscience Institute, University of Iowa, Iowa City, IA

## Abstract

Language abilities are highly heritable, yet their genetic architecture remains poorly understood due to limitations in scalable phenotyping. Traditional clinical assessments are time-intensive (often 1-3 hours) and require specialized personnel, fundamentally constraining the sample sizes necessary for robust genetic discovery. Here we introduce Lingo, an open-source web-based platform that captures comprehensive language and cognitive phenotypes in approximately 30 minutes through seven tasks administered remotely. Critically, comparative power analyses demonstrate that Lingo-derived phenotypes achieve nearly 2-fold greater statistical power than clinical questionnaire measures and *>*10-fold improvement over self-reported language diagnoses for detecting polygenic score associations. This translates to meaningful sample size reductions: 1,000 participants using Lingo is equivalent to 1,730 with in-depth questionnaires or *>* 13,000 people with self-reported language impairment. Analyzing Lingo data from *>* 2,000 adults, we identify four interpretable factors with high test-retest reliability (*r* = 0.69–0.79): narrative fluency, reading fluency, phonemic fluency, and general cognitive ability (*g*). The *g* factor demonstrates strong concurrent validity with clinical Wechsler full-scale IQ assessment (*r* = 0.78). These factors reveal distinct psychiatric and genetic profiles: *g* associates with externalizing behaviors and ADHD polygenic propensity, while phonemic fluency links specifically to withdrawn behavior as well as depression and schizophrenia polygenic scores. Rare variant burden analysis of Lingo scores identifies novel candidate genes (*NGB* and *GLS*) and implicates ATP metabolism and white matter pathways in language ability. These results establish Lingo as a transformative tool that makes previously impractical genetic studies feasible by enabling adequately powered studies with dramatically reduced sample sizes and costs.

## Introduction

Language represents one of humanity’s most complex cognitive abilities, integrating multiple brain networks to enable communication, thought, and social interaction [1]. Individual differences in language ability are substantial and highly heritable, with twin studies often demonstrating heritability estimates of 45-65% for various language phenotypes [2–4]. However, despite this strong genetic basis, the genetic architecture of language abilities remains poorly understood. Genome-wide association studies (GWAS) have primarily focused on binary traits (e.g., dyslexia [5], stuttering [6], and rhythm [7, 8]) or proxy measures (e.g., educational attainment [9]), with few large-scale efforts examining quantitative language abilities directly [10, 11]. For the quantitative language traits that have been studied, progress has been limited by the challenge of achieving the large sample sizes necessary for dissecting highly polygenic traits. This challenge is exacerbated by methodological constraints in phenotyping that make large-scale data collection particularly difficult for language abilities.

Traditional clinical language assessments present significant barriers to large-scale research: they are time-intensive (often 1-3 hours per participant) and require specialized personnel to administer. Consequently, even the largest genetic studies of quantitative language-related traits struggle to achieve sample sizes comparable to other cognitive domains (N *≈* 34,000 for reading traits versus N *>* 250,000 for other cognitive domains) [10, 12]. The recent GenLang consortium meta-analysis of reading traits in over 30,000 participants highlighted these limitations [10], calling for innovative phenotyping tools to enable the sample sizes necessary for genetic discovery. Beyond simply reducing time and related costs, achieving these sample sizes requires measurement approaches that maximize statistical power per participant. Phenotypes with reduced measurement error and greater sensitivity to genetic variation can achieve equivalent statistical power with substantially smaller samples, fundamentally changing the feasibility calculus for genetic discovery studies.

Language phenotyping is equally important for psychiatric and neurodevelopmental research, where language skills serve as both risk markers and diagnostic features. Children with language impairments face 2-3 fold increased psychiatric risk later in life [13, 14], while distinct language signatures emerge across psychiatric conditions: reduced semantic coherence in schizophrenia [15–17], altered emotional prosody in depression [18], and high comorbidity with ADHD and autism [19, 20]. Language markers hold promise for improving diagnostic precision in psychiatry, yet their systematic study faces the same methodological barriers limiting genetic research.

Recent work has shown promising results that online assessments can accurately measure individual differences in cognitive and language abilities and may even be feasible for genetic research. One assessment, Pathfinder, is a 15-minute online gamified measure of general cognitive ability (*g*), has demonstrated brief web-based tests can capture substantial genetic variance, with twin heritability estimates of 57% and polygenic scores predicting up to 12% of variance in cognitive performance [21]. Similarly, the Individual Differences in Language Skills (IDLaS-NL) test battery, a set of 31 assessments administered online in Dutch, has shown strong construct validity for measuring variability in spoken language production, comprehension, linguistic knowledge, and related cognitive skills in adults [22–24]. While these developments demonstrate the feasibility of online assessment, neither approach simultaneously achieves the comprehensive phenotyping, genetic validation, and statistical efficiency needed to make large-scale genetic studies of language practical. Pathfinder provides genetic validation but lacks deep language phenotyping, while IDLaS-NL offers comprehensive language assessment without demonstrated genetic utility. This creates a critical gap: no existing tool combines scalable administration, fine-grained multidimensional language phenotyping, and the statistical power necessary for genetic discovery studies with realistic sample sizes.

To address these challenges, we developed Lingo, an open-source comprehensive web-based language battery administered remotely in approximately 30 minutes. By democratizing access to comprehensive language assessment through freely available tools that require no specialized training or expensive equipment, Lingo removes traditional barriers that have limited growth in the field of language genetics. Lingo assesses a variety of language and cognitive skills, including: expressive language, receptive language, rhythm, reading, and nonverbal abilities. Lingo captures multiple data streams, including: item selections, response time, and high-quality audio recordings that enable analysis of acoustic, linguistic, and cognitive phenotypes. Here, we validate Lingo across over 2,000 individuals, demonstrating high test-retest reliability and significant correlations with established cognitive assessments, and present analyses revealing distinct genetic and psychiatric profiles of language phenotypes. Notably, comparative power analysis shows Lingo-derived phenotypes achieve *≈*2-fold greater statistical power for polygenic score associations compared to traditional questionnaire measures, and *>*10-fold greater power versus self-reported dyslexia and language impairments, establishing Lingo as a transformative tool that makes previously impractical genetic studies feasible for individual research groups with dramatically reduced sample sizes and costs.

## Results

### Lingo overview

An overview of the Lingo battery and this study can be seen in Figure 1. Prompt images can be seen in the Supplementary Figures and descriptions of each task can be found in the “Methods” section. Open source code of Lingo is available at https://research-git.uiowa.edu/michaelson-lab-public/lingo. For convenience, we also provide a pre-compiled executable tool so that other researchers can easily use Lingo for data collection.

**Figure 1.**
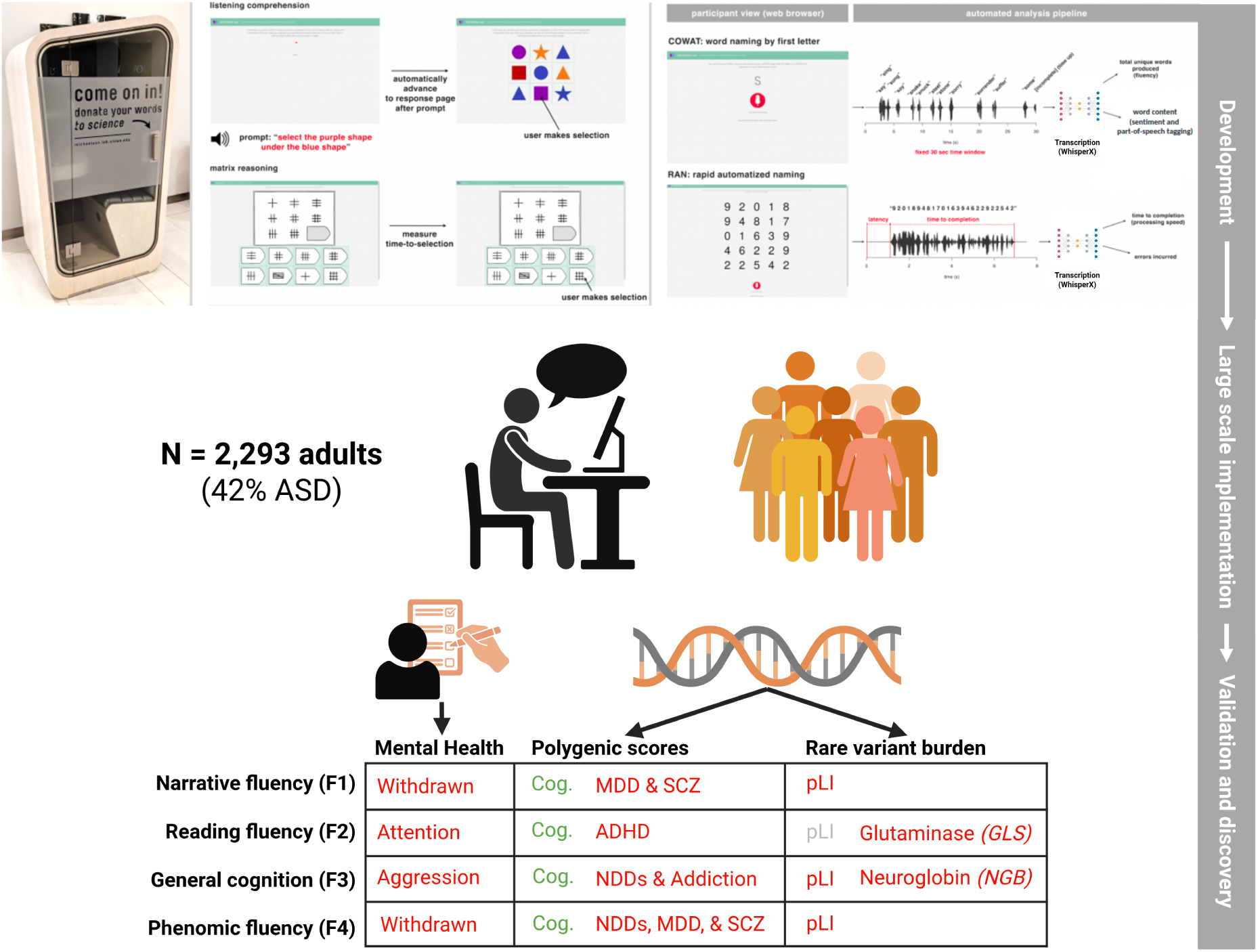
Graphical overview of Lingo and this study. Lingo consists of seven tasks aimed at measuring individual differences in language and cognition. Raw audio, selections, and timing are recorded during Lingo. Representative screenshots from Lingo tasks are shown here as well as examples of the data produced. Additional details about data collection in SPARK, validation, and a table summarizing key findings are shown here. Red text in the results summary table indicate negative correlations (e.g., higher Narrative Fluency scores on Lingo were associated with being less withdrawn), green text indicates positive correlations (e.g., higher Narrative Fluency scores on Lingo were associated with higher cognitive polygenic scores), and grey indicates no correlation. Cog. = cognitive performance, MDD = major depressive disorder, SCZ = schizophrenia, ADHD = Attention-Deficit/Hyperactivity Disorder, NDDs = neurodevelopmental disorders, pLI = probability of a gene being loss-of-function intolerant. Figure was created with BioRender.

### Lingo performance features and their factor structure

The open-ended nature of many Lingo screener tasks provides flexibility in measuring performance, enabling both hypothesis-driven and exploratory analyses. We extracted 41 item-level features from Lingo tasks that passed quality control (ranging from accuracy scores to response times to word counts and others; see supplementary information Table S1 for detailed descriptions), which we use throughout this manuscript to measure different aspects of language and cognitive abilities. These features capture performance metrics related to verbal memory, verbal fluency, reading speed, receptive language, rhythm, and nonverbal abilities. We hypothesized that these features might reflect one or more shared underlying constructs.

To investigate this, we conducted an exploratory factor analysis, which identifies groups of correlated variables that measure common underlying abilities. The factor analysis was performed on a large sample recruited through the SPARK Research Match platform, a genetically informed autism cohort [25, 26]. Our sample included 2,293 adults (mean age = 40.2 years; 1,722 females, 571 males), of whom 962 had autism (591 females, 371 males).

The Lingo data was suitable for factor analysis, as indicated by Bartlett’s sphericity test (p *<* 2*×*10*^−^*^16^) and the Kaiser-Meyer-Olkin criterion (KMO = 0.88). Using a data-driven approach (see Methods), we identified four factors from our Lingo features (Figure 2a). Factor F1 (narrative fluency) loaded primarily onto the number of words produced during the Picture Narration task, reflecting spontaneous expressive language generation. Factor F2 (reading fluency or naming fluency) loaded strongly onto completion speed for the Rapid Automatized Naming (RAN) tasks, capturing rapid naming abilities that are strongly associated with reading and dyslexia [27]. Factor F3 showed strong loadings across sentence repetition, matrix reasoning, rhythm, and following directions tasks. We interpret this factor as likely reflecting general language ability, given the linguistic demands common to most of these tasks, though it’s loading onto matrix reasoning suggests overlap with general cognition. We refer to it as the *g* factor given the extensive prior literature on this construct. Notably, indices related to receptive language also loaded onto this factor. Factor F4 (phonemic fluency) loaded onto the number of words produced during the COWAT task, measuring lexical retrieval and executive control. Loadings for all factors and features can be found in Table S2. Factor score inter-correlations are shown in Figure 2b, with the phonemic fluency factor (F4) showing the strongest correlations across all factors.

**Figure 2.**
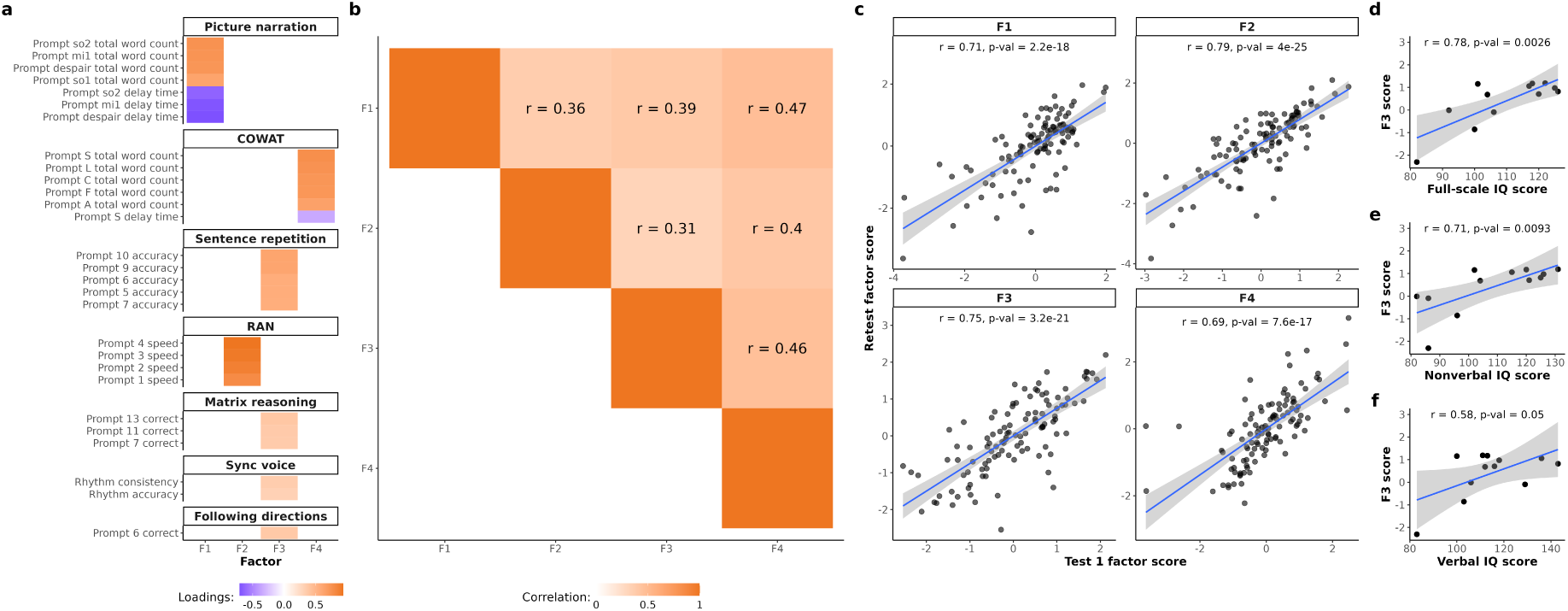
The factor structure and psychometric properties of Lingo. **a** Factor loadings of Lingo features (only items with a *|loading| >* 0.3 are shown). F1 = narrative fluency, F2 = reading fluency, F3 = *g* (general cognitive ability), F4 = phonemic fluency. Orange values indicate positive loadings, while purple indicate negative loadings. **b** Correlation heatmap of Lingo factors. Orange indicates positive correlations. Pearson correlation coefficients are shown (*r*). **c** Correlation between Lingo factor scores for individuals who took the assessment twice (N = 111), demonstrating high stability across a 2+ week interval. X-axis is the factor score from the first assessment, Y-axis is the factor score from the second assessment (taken at least 2 weeks later). Pearson correlation statistics are shown. **d, e, f** Correlations between clinical Wechsler IQ scores and Lingo F3 scores (N = 12 individuals). Pearson correlation statistics are shown.

### Lingo shows substantial test-retest reliability

To investigate test-retest reliability of Lingo scores, we asked 111 individuals from the SPARK research match study to take the Lingo battery twice (separated by at least two weeks between tests). Test-retest reliability was measured by calculating Pearson correlations and intraclass correlation coefficients (ICC) between the factor scores from the first and second testing sessions, on a factor-wise basis (Figure 2c). The factor model was fit using data from all first testing sessions, and this model was then used to predict factor scores for the second sessions (see Methods). All factors had statistically significant test-retest reliability correlations. F1-F3 showed high test-retest reliability (Pearson *r* = 0.71–0.79, Supp Table S3). F4 (phonemic fluency), showed reasonable test-retest reliability (Pearson *r* = 0.69, p = 7.6 *×* 10*^−^*^17^) given that phonemic fluency tasks are known to exhibit practice effects [28].

All factors showed lower ICC scores than Pearson correlation coefficients, suggesting a small but consistent shift in factor scores across sessions (for example, F3 *r* = 0.75, F3 ICC = 0.72, Table S4). We found this is likely a learning effect, as individuals tended to score higher across Lingo factors the second time they take it (Figure S3a; Table S5). Notably, we show individual differences in this learning effect could be an interesting trait to study, with learning magnitude correlating with polygenic scores (see “Polygenic associations of Lingo reveal genetic overlap between language and psychiatric conditions” section).

### Lingo scores correlate with established cognitive assessments

To assess the concurrent validity of the Lingo battery, we examined the relationship between factor scores and standardized measures of cognitive ability. Specifically, we compared our Lingo factor scores to clinical Wechsler Intelligence Scale assessments, which are considered gold-standard measures of general intelligence. A subset of twelve individuals from our SPARK sample had complete Wechsler IQ data available.

We found that the factor interpreted as general intelligence (*g* ; F3) was significantly correlated with Full Scale IQ (*r* = 0.78, p = 0.003), Nonverbal IQ (*r* = 0.71, p = 0.009), and Verbal IQ (*r* = 0.58, p = 0.05, Figure 2d). Despite this modest validation sample size, these correlations support the interpretation that F3 represents general cognitive ability (*g*). The strength of these correlations (*r* = 0.58–0.78) is comparable to or exceeds typical convergent validity coefficients between different IQ tests [29], suggesting that Lingo provides an efficient proxy for comprehensive cognitive assessments. No other factor was significantly associated with the IQ assessments (Table S7).

### Mental health associations with Lingo scores reveal psychiatric comorbidities

To examine whether Lingo-derived language phenotypes may capture clinically relevant information for psychiatric assessment, we assessed their associations with mental health symptoms using the Adult Self-Report questionnaire (ASR, N = 1,514 with Lingo and ASR scores). In order to identify specific symptom domain associations beyond general psychiatric/behavioral problems, we included the Total Problems T-Score (as well as age and sex) as covariates in all analyses of subscales. This approach revealed distinct patterns of association between language factors and psychiatric symptomatology (Figure 3a, full summary statistics can be found in Table S7).

**Figure 3.**
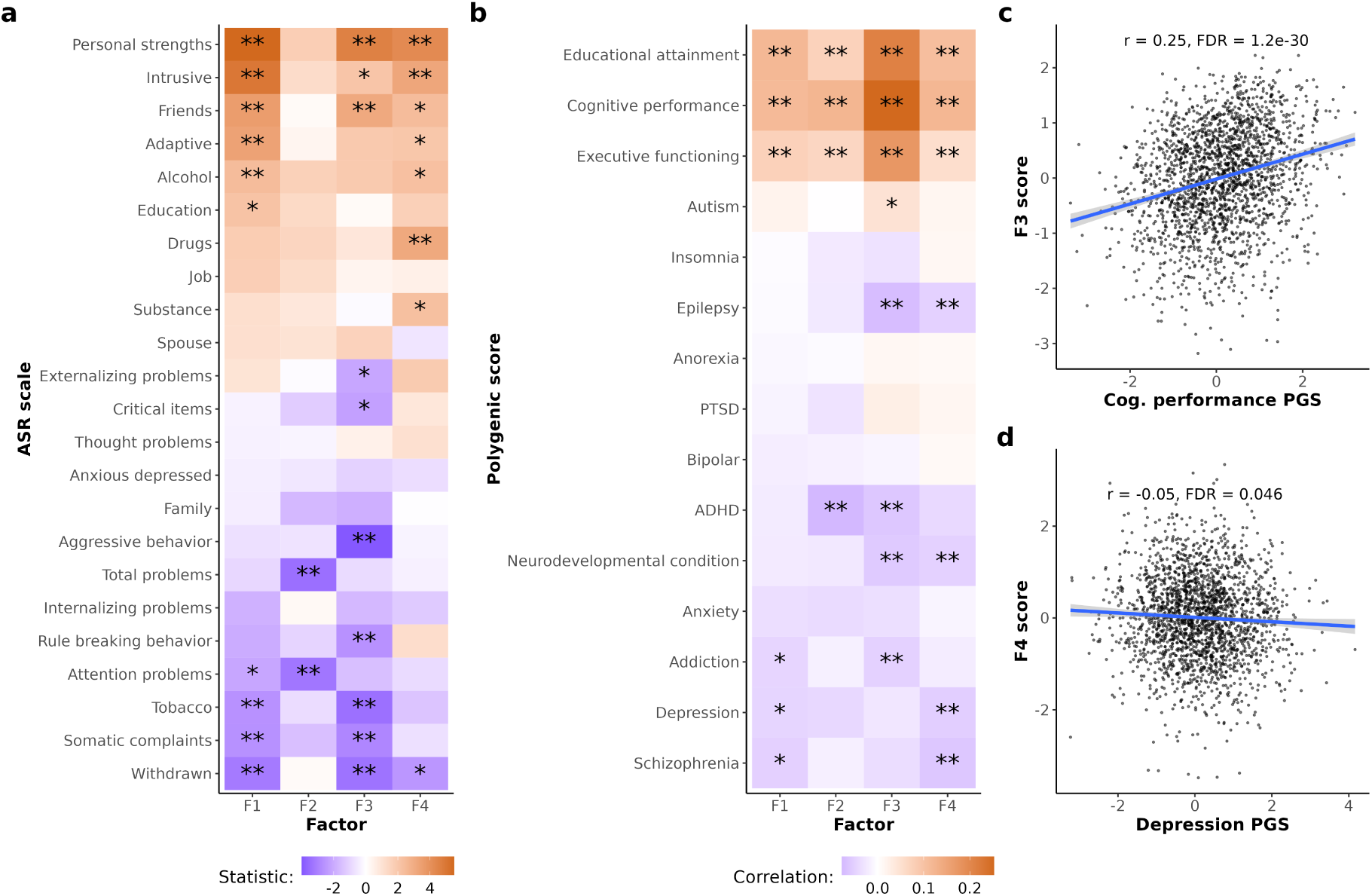
Lingo factors exhibit differential associations with mental health, behaviors, and polygenic scores. (**a**) Heatmap showing associations between Lingo factors (F1-F4) and Adult Self-Report (ASR) scale measures, calculated using linear regression models (N = 1,514). Color intensity indicates effect size; orange = positive associations, purple = negative associations. Significance levels: ** FDR-adjusted p *<* 0.05, * p *<* 0.05. (**b**) Correlations between Lingo factors and polygenic scores (PGS) for various psychiatric and cognitive traits (N = 2,098). Color intensity indicates effect size; orange = positive associations, purple = negative associations. (**c**) Scatter plot showing correlation between cognitive performance PGS (X-axis) and *g* factor scores (F3). Pearson correlation statistics are shown. (**d**) Scatter plot showing correlation between depression PGS (X-axis) and phonemic fluency factor scores (F4). Pearson correlation statistics are shown.

Narrative fluency (F1) showed extensive FDR-significant psychiatric symptom associations, including positive associations with personal strengths (*β* = 0.13, FDR = 1.1 *×* 10^-6^), intrusive behaviors (*β* = 0.11, FDR = 9.2 *×* 10^-6^), friends (*β* = 0.09, FDR = 0.003), adaptive functioning (*β* = 0.07, FDR = 0.005), and alcohol use (*β* = 0.06, FDR = 0.046). Narrative fluency also showed negative associations with withdrawn behavior (*β* = -0.05, FDR = 0.008, tobacco use (*β* = -0.07, FDR = 0.031) and somatic complaints (*β* = -0.05, FDR = 0.031). This pattern indicates that higher narrative fluency as ascertained by Lingo relates to better social functioning and adaptive skills.

Reading fluency (F2) demonstrated the fewest links with mental health symptoms, showing only negative association with total problems (*β* = -0.08, FDR = 0.013) and attention problems (*β* = -0.05, FDR = 0.013). This limited pattern suggests that reading fluency measured in Lingo primarily relates to general mental health problems and attention issues rather than specific symptom clusters seen with other Lingo factor scores.

*g* factor scores (F3) showed robust associations with specific domains, including positive correlations with personal strengths (*β* = 0.11, FDR = 1 *×* 10^-4^) and friends (*β* = 0.08, FDR = 0.009). *g* scores showed negative associations with aggressive behavior (*β* = -0.07, FDR = 9 *×* 10^-4^), tobacco use (*β* = -0.09, FDR = 0.005), withdrawn behavior (*β* = -0.06, FDR = 0.005), somatic complaints (*β* = -0.05, FDR = 0.016), and rule-breaking behavior (*β* = -0.05, FDR = 0.03). This pattern aligns with previous work finding that higher cognitive ability is associated with fewer mental health problems, particularly with externalizing behaviors.

Phonemic fluency scores (F4) exhibited relatively few FDR-significant associations, showing positive associations with personal strengths (*β* = 0.1, FDR = 5.1 *×* 10^-4^), drug use (*β* = 0.08, FDR = 0.014), intrusive behaviors (*β* = 0.07, FDR = 0.014). Withdrawn behavior was the only variable that showed a negative association with this factor, and this association was nominal (*β* = -0.04, FDR = 0.075). This suggests that higher phonemic fluency, as formulated by this factor analysis, may not be as predictive of mental health problems compared to the other Lingo factors.

Taken together, these results suggest that Lingo captures language traits relevant to mental health. While higher performance on any Lingo factor appears to generally be associated with fewer mental health related problems, there is specificity in symptom domains across Lingo factors. For example, higher *g* (F3) scores were associated with fewer externalizing symptoms, while higher reading speed (F2) was linked to less attention problems. The differential patterns offers preliminary support for the utility of Lingo in psychiatric phenotyping.

### Polygenic associations of Lingo reveal genetic overlap between language and psychiatric conditions

To examine the genetic architecture underlying Lingo-derived language phenotypes, we analyzed associations between Lingo factor scores and polygenic scores (PGS) for cognitive and psychiatric traits (N = 15 PGS spanning cognitive and psychiatric traits, N = 2,098 individuals with both Lingo and PGS data). This analysis revealed genetic profiles across language components (Figures 3b-d).

All four Lingo factors showed robust associations with cognitive-related polygenic scores. Cognitive performance, educational attainment, and executive functioning PGS were significantly positively associated with all four Lingo factors, indicating shared genetic architecture between language and cognitive abilities measured by Lingo and established genetic factors influencing cognitive functioning. Notably, *g* (F3) demonstrated the strongest association with cognitive performance PGS (*r* = 0.25, FDR = 1.2 *×* 10^-30^; Figure 3c), consistent with its role as a general cognitive ability factor.

Psychiatric polygenic scores revealed more selective association patterns, with weaker effects than cognitive polygenic scores. Epilepsy PGS showed significant negative associations with *g* (F3, *r* = -0.07, FDR = 0.004) and phonemic fluency (F4, *r* = -0.05, FDR = 0.049). ADHD PGS was negatively associated with both reading fluency (F2, *r* = -0.08, FDR = 0.003) and *g* (F3, *r* = -0.06, FDR = 0.016). Rare neurodevelopmental condition PGS demonstrated negative associations with *g* scores (F3, *r* = -0.06, FDR = 0.021) and phonemic fluency (F4, *r* = -0.05, FDR = 0.049). Depression PGS showed a specific negative association with phonemic fluency (F4, *r* = -0.05, FDR = 0.046; Figure 3d), similarly schizophrenia PGS was also negatively associated with phonemic fluency (F4, *r* = -0.06, FDR = 0.027). Additionally, addiction PGS showed a negative association with *g* (F3; *r* = -0.05, FDR = 0.048). Autism, insomnia, anorexia, PTSD, bipolar, and anxiety PGSs had no significant associations after multiple testing correction. Full summary statistics for the correlations between PGS and Lingo factors can be found in Table S8.

These findings reveal that while all Lingo factors share genetic overlap with cognitive abilities, psychiatric genetic loci load differentially across language phenotypes. *g* (F3) and phonemic fluency (F4) showed the most psychiatric associations, suggesting they may be particularly sensitive to genetic variants influencing mental health, while narrative fluency (F1) showed no FDR significant associations with psychiatric PGS in our sample. The selective pattern of associations supports the genetic distinctiveness of different language components and their differential relationships with cognitive versus psychiatric genetic architectures.

In a subset of participants who completed Lingo twice (N = 109 with PGS data and multiple Lingo assessment scores), we observed systematic improvements across all language factors between sessions (likely a weak learning effect, Figure S3a), suggesting Lingo can capture individual differences in ability to learn language. Notably, the magnitude of improvement (difference between Lingo Test 2 and Test 1 scores) correlates with polygenic scores: individuals with higher ADHD PGS demonstrated less improvement (*r* = -0.31, p = 9.8 *×* 10^-4^; Figure S3b), while those with higher cognitive performance PGS showed a trend toward greater improvement (*r* = 0.18, p = 0.069; Figure S3c). These preliminary findings suggest that Lingo may capture not only static language and cognitive abilities but also dynamic learning processes, with polygenic scores influencing individual capacity for language skill acquisition.

### Rare variant associations of Lingo converge on neurometabolism and white matter pathways

Next, to show the utility of Lingo for discovery of specific genes and biological pathways underlying language abilities, we performed rare variant association analyses using whole-exome sequencing data (N = 1,671 with Lingo and WES data). As a proof of principle, we hypothesized that damaging variants in mutation-intolerant genes, which are robustly linked to cognition and neurodevelopment [30–33], would be associated with lower Lingo scores. To test this hypothesis, we examined the cumulative burden of rare damaging variants (predicted loss of function or missense variants with a minor allele frequency *<* 1%) in genes with a pLI score *>* 0.9 and determined whether this rare genetic burden score was predictive of Lingo factor scores (including age, sex, ancestral background, and genome wide burden as covariates). As expected, we found that individuals carrying more rare damaging variants in mutation-intolerant genes showed significantly lower *g* factor scores (F3, *β* = -0.13, p = 4.3 *×* 10*^−^*^4^; Figure 4a), with similar negative associations observed for narrative fluency (F1) and phonemic fluency (F4, Table S9). This suggests that rare genetic variants in genes under selective constraints have detrimental effects on cognitive and language abilities measured by Lingo.

**Figure 4.**
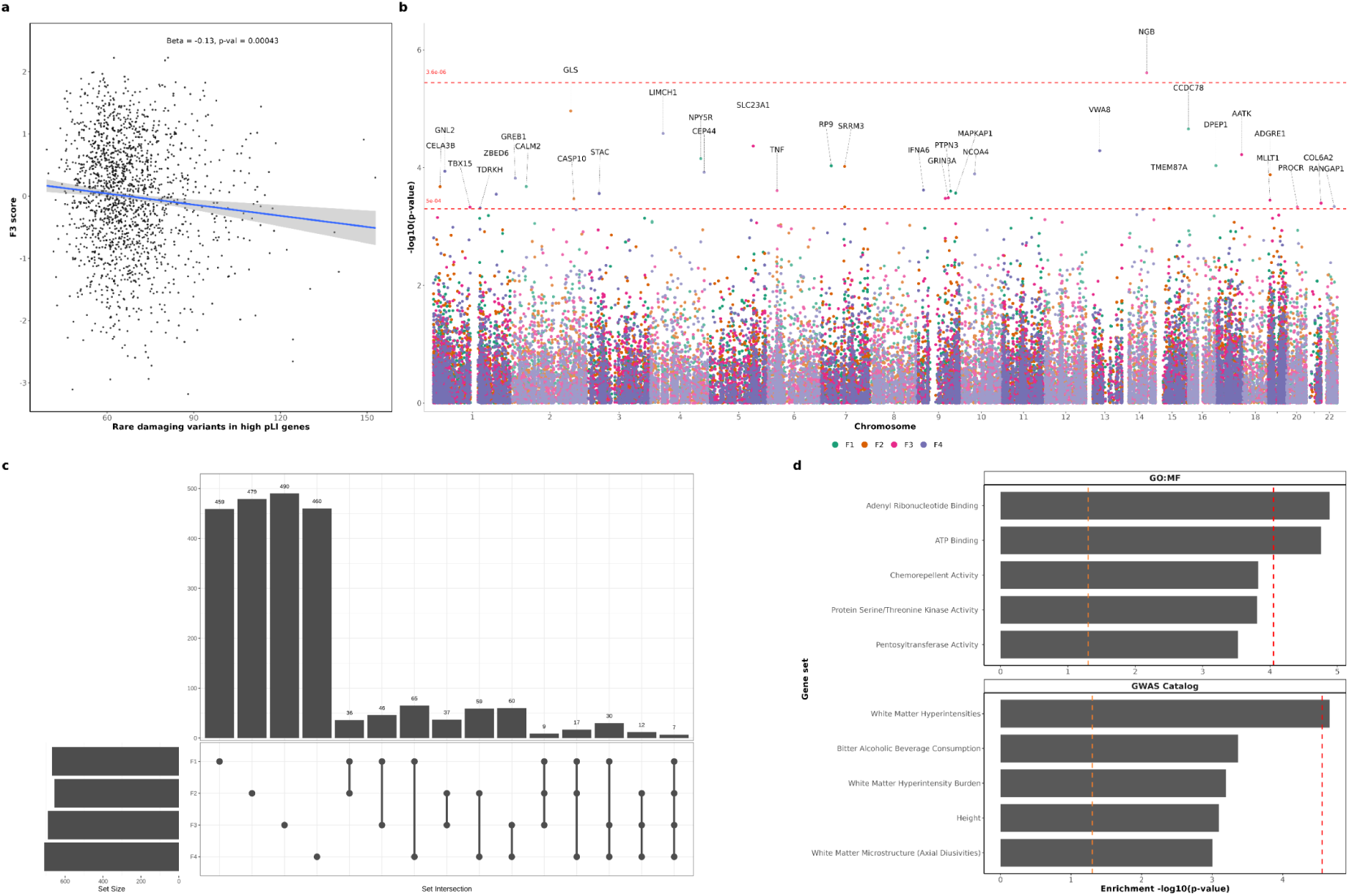
Rare variant analysis reveals novel candidate genes and biological pathways associated with language abilities. (**a**) Association between rare damaging variants in high pLI (X-axis; probability of loss-of-intolerance) genes and *g* (F3) factor scores (Y-axis). Linear regression statistics are shown (N = 1,671). Lines showing the genome-wide significant and suggestive cutoffs are shown and genes passing theses thresholds are labeled. (**b**) Manhattan plot showing gene-level genome-wide association results across Lingo factors. (**c**) Upset plot showing gene-level association overlap across factors; horizontal bars show total genes associated with each factor, vertical bars show genes shared between factor combinations. (**d**) Gene Ontology and GWAS catalog enrichment analysis of all nominally associated genes with any Lingo factor score. Red dotted line indicates Bonferroni statistical significance cutoff, and orange indicates nominal statistical significance.

Next, we conducted gene-level association testing (see Methods), which revealed neuroglobin (*NGB*), a protein involved in neuronal oxygen transport and protection against oxidative stress, as a novel candidate gene for general cognitive ability (*g*, F3; p = 2.4 *×* 10*^−^*^6^, Bonferroni-corrected p = 0.034; Figure 4b). Additionally, glutaminase (*GLS*), a key enzyme in glutamate neurotransmission, showed a suggestive association with reading fluency (F2; p = 1.1 *×* 10*^−^*^5^, Bonferroni-corrected p = 0.15), implicating excitatory neurotransmission in rapid automatized naming. Examining the gene-level results of large GWASs provides independent validation for the effect of these genes on language and cognitive traits (see Methods). Both *NGB* and *GLS* have been implicated in the most recent educational attainment GWAS (MAGMA p = 0.002 and 0.018, respectively), and *GLS* was previously implicated in the largest dyslexia GWAS (MAGMA p = 5.6 *×* 10*^−^*^4^) [5, 9].

Investigating genetic overlap across factors demonstrated that most gene associations were factor-specific (nominal p *<* 0.05, (Figure 4c), suggesting distinct rare variant architectures underlying different aspects of language ability. Narrative fluency (F1) and phonemic fluency (F4) showed the most overlap across factors (65 genes). Gene set enrichment analysis of all language-associated genes (any gene with nominal p *<* 0.05) revealed significant enrichment for several biological processes (Figure 4d). Molecular function analyses highlighted ATP binding and adenyl ribonucleotide binding pathways, implicating cellular energy metabolism in language abilities. Analysis of GWAS catalog traits [34] showed enrichment for genes previously associated with white matter hyperintensities. These rare variant findings complement our polygenic score analyses by identifying specific genes and biological pathways contributing to language variation. The convergence on ATP binding processes and white matter integrity suggests that language abilities depend on efficient neural energy metabolism and structural brain connectivity. The identification of novel candidate genes like *NGB* and *GLS* provides new targets for understanding the molecular mechanisms underlying language-related disorders.

### Lingo improves statistical power for genetic associations

Finally, to determine whether Lingo may provide greater power for detecting genetic associations than more commonly used measures like self-report data, we conducted a comparative power analysis. We used the cognitive performance PGS as a benchmark, given its established associations with language-related traits [5, 10, 35]. We compared Lingo *g* (F3) against more traditional measures of language available in the SPARK sample. These measures included the Communication Standard Score from the VABS-3 questionnaire (an in-depth measure often used to support clinical diagnosis of intellectual and developmental disabilities [36]) and self-reported language-related disorders (dyslexia and language impairment). Power was defined as the ability to detect significant associations between the phenotype of interest (e.g., Lingo F3 score) and the cognitive performance PGS across varying sample sizes (100-1,500 participants, with stepwise increases of 25 participants), with 1,000 bootstrap samples at each sample size. Age, sex, and genetic ancestry were included as covariates.

Lingo demonstrated markedly superior statistical power compared to conventional measures (Figure 5a). Lingo achieved 80% power to detect statistically significant (p *<* 0.05) cognitive performance PGS associations with 122 individuals, while VABS-3 required approximately 209 to reach the same threshold. Self-reported language disorders showed substantially lower power, with neither language impairment or dyslexia achieving 80% power to detect cognitive performance PGS associations, despite known genetic correlations between cognitive performance and dyslexia and quantitative reading traits [5, 10]. To achieve 80% power to detect cognitive performance PGS associations we approximate 1,627 samples self-reporting dyslexia would be needed, and 1,877 samples for language impairment would be needed. It is important to note that the self-report power analysis is likely sensitive to case-control imbalances, our sample had *>* 7% self-reporting language impairment and/or dyslexia, which is a slightly higher rate compared to the largest dyslexia GWAS to date (4.5%) [5].

**Figure 5.**
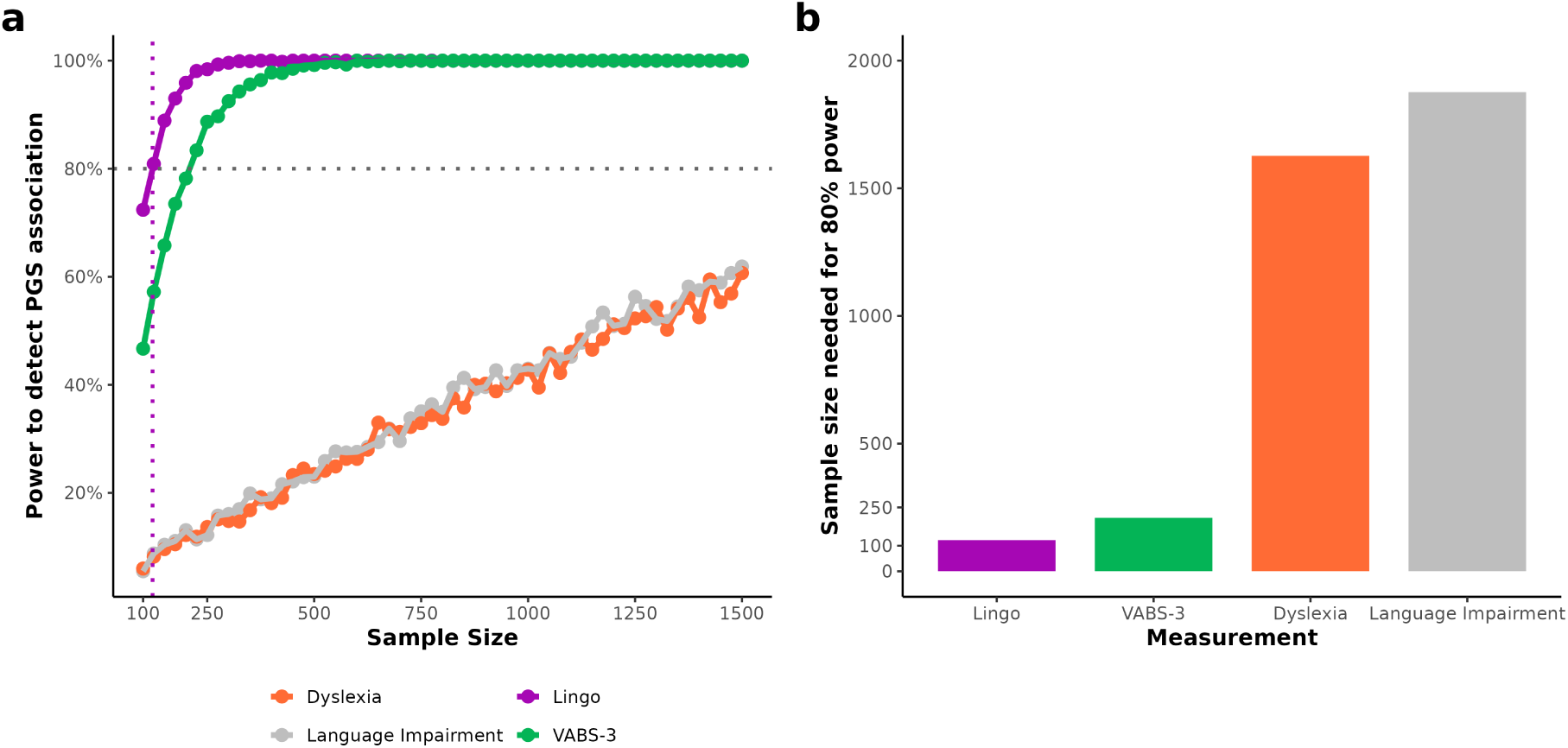
Lingo demonstrates superior statistical power for genetic analysis compared to traditional language assessments. (**a**) Power analysis showing detection rates for cognitive performance polygenic score (PGS) across language phenotyping methods. X-axis indicates sample size, Y-axis is power to detect an association (proportion of bootstrap samples with statistically significant association). Dotted horizontal line indicates 80% power threshold. (**b**) Comparison of estimated sample size needed to for 80% power to detect associations with the cognitive performance PGS across language phenotyping methods.

Comparing the number of samples needed to reach 80% power to detect cognitive performance PGS associations reveals dramatic differences in efficiency between methods (Figure 5b). Our data indicates that Lingo reduces the number of samples needed to reach 80% power by 41.6% compared to the VABS-3. Self-reported measures showed markedly reduced efficiency, with Lingo needing 92.5% fewer samples than dyslexia diagnosis and 93.5% fewer samples than language impairment diagnosis.

These findings demonstrate that Lingo-derived phenotypes can provide substantial advantages for genetic discovery studies. The improved power translates to reduced sample size requirements which should lead to substantially increased cost-effectiveness for research investigating the genetic architecture of language abilities. The observed power increase likely reflects Lingo’s comprehensive assessment of multiple language domains and reduced measurement error compared to questionnaires or binary self-report measures.

## Discussion

Traditional language assessments present significant barriers to genetic discovery: requiring specialized personnel, controlled environments, and 1-3 hours to assess each participant. These challenges fundamentally limit achievable sample sizes to far less than the N *>* 100,000 typically necessary for genetic discovery. We developed Lingo to address this bottleneck through scalable digital phenotyping that maintains clinical validity while dramatically improving statistical efficiency. Analyzing data from *>* 2,000 individuals, we demonstrate that Lingo-derived phenotypes provide roughly 2-fold greater power than clinically validated questionnaires and *>* 10-fold improvement over self-reported language diagnoses for detecting polygenic associations. With Lingo, we identified four language factors with distinct psychiatric and genetic profiles, discovered novel candidate genes (*NGB*, *GLS*), and implicated ATP metabolism and white matter pathways in language and cognitive phenotypes. These findings demonstrate that digital language phenotypes capture biologically meaningful variation relevant to psychiatric research.

The statistical advantages of Lingo can make genetic studies of language more feasible. Our power analyses demonstrate that detecting a polygenic association requires 122 participants with Lingo, 209 participants with clinically relevant questionnaires, or *>* 1,500 participants using self-reported diagnoses. Extrapolating to genome-wide association study requirements (N*>*100,000), questionnaire-based studies would need *>* 170,000 participants while diagnostic (i.e., case/control) approaches would require over one million participants to achieve equivalent power to 100,000 Lingo participants. This efficiency gain stems from the capture of objectively measured language abilities rather than relying on self-reported traits.

Beyond statistical power advantages, Lingo also removes many of the practical barriers that have long limited large-scale research on language ability. Conducting a reasonably well powered study for polygenic score discovery using standard in-person language assessments (N *≈* 1,000) could cost 1,000–3,000 clinician hours, and take months or years of data collection. In contrast, data collection with Lingo incurs minimal costs beyond participant compensation, requires no specialized personnel, and the online nature of Lingo means it can be completed in parallel at anytime by participants leading to faster data collection. This represents a significant reduction in time and potential costs, greatly expanding the feasibility of adequately powered studies. Moreover, Lingo’s remote administration enables global collaboration without site visits, frequent longitudinal follow-ups with minimal participant burden, and could increase research participation by under-resourced institutions.

Beyond practical and methodological advantages, our psychiatric and genetic analyses reveal that Lingo factors capture biologically meaningful variation with relevance to mental health. Phonemic fluency (F4) demonstrated striking convergence across independent sources of information: nominal negative associations with self-reported withdrawn behavior alongside FDR-significant negative correlations with both depression and schizophrenia polygenic scores. Given that social withdrawal represents a core clinical feature of both diagnoses [37, 38], this convergence suggests that reduced phonemic fluency may capture shared neurobiological mechanisms underlying social disengagement across psychiatric conditions. The general cognitive factor (F3) showed complementary patterns with externalizing psychopathology, supporting previous work that has shown higher *g* scores correlate with reduced aggressive behavior and rule-breaking behavior [39, 40], alongside negative associations with ADHD and addiction polygenic scores. These factor-specific patterns support distinct biological architectures underlying different language components and suggest potential applications as differential markers across psychiatric conditions.

Our rare variant analyses demonstrate that Lingo can enable gene discovery with samples far smaller than typical case/control GWAS requirements, providing biological insights into language abilities. Neuroglobin (*NGB*) emerged as a genome-wide significant candidate gene for general cognitive ability. *NGB* ’s role in neuronal oxygen homeostasis and neuroprotection [41] suggests that cellular stress response mechanisms contribute to cognitive variation in the general population, extending previous findings in neurodevelopment [42, 43]. The replication in educational attainment GWAS (MAGMA p = 0.002) strengthens confidence in this association [9]. The association between rare variants in glutaminase (*GLS*) and reading fluency implicates glutamatergic neurotransmission in rapid automatized naming. *GLS* converts glutamine to glutamate, the brain’s primary excitatory neurotransmitter, connecting to neuroimaging evidence of altered glutamate signaling in dyslexia [44, 45] and suggesting potential pharmacological targets. Critically, *GLS* shows convergent evidence in the dyslexia GWAS (MAGMA p = 5.6 *×* 10*^−^*^4^), providing orthogonal validation [5]. The pathway-level enrichment for ATP metabolism and white matter integrity converges with genetic and neuroimaging evidence that reading fluency depends on efficient white matter connectivity and metabolic support for rapid information processing [46–48]. This molecular-to-systems convergence exemplifies how scalable phenotyping can bridge genetic variants to brain biology.

Several limitations suggest priorities for future development. First, while Lingo’s web-deployed version requires internet access, we provide a containerized version that can be hosted locally (e.g., on a laptop) without ongoing internet connectivity. However, both deployment options require a computer with a microphone and digital literacy, which may limit accessibility or introduce sampling bias in some populations. Second, Lingo is currently implemented and validated in English-only. Cross-linguistic validation is needed, though many tasks (Matrix Reasoning, Sync Voice, RAN) require minimal language-specific modification, and we welcome collaborations for adaptation. Additionally, Lingo has been validated only against Wechsler IQ assessments and polygenic estimates of general cognitive abilities; further validation with clinical language assessments such as the CELF would increase confidence that Lingo captures distinct aspects of language ability beyond general cognitive skills. Third, we validated Lingo in adults (18+); extension to children may require age-appropriate modifications and separate validation. Fourth, while our sample included individuals with autism, broader clinical validation (e.g., aphasia, primary language disorders, schizophrenia) is needed before clinical deployment, though 58% of our participants did not have autism and we demonstrate robust genetic associations with general population traits. Fifth, Lingo does not capture information on all aspects of language. For example, Lingo does not currently include any written language or grammar-focused tasks and consequently cannot capture deficits specific to these areas.

In summary, Lingo is a powerful digital platform for phenotyping language and cognition that simultaneously demonstrates clinical validity, psychiatric relevance, and genetic associations, while offering 2–10-fold gains in statistical power over conventional assessments. This efficiency could transform research feasibility, making adequately powered language genetics studies accessible to individual research groups. The convergent psychiatric and genetic findings provide new insights into the biological basis of language, and the identification of novel candidate genes (*NGB*, *GLS*) together with the involvement of ATP metabolism and white matter pathways highlight potential molecular mechanisms underlying individual differences in language ability. As digital phenotyping continues to advance, platforms like Lingo are poised to play a central role in precision psychiatry and in uncovering the genetic foundations of human cognition.

## Methods

### Lingo Platform Architecture and Implementation

Lingo is a custom-built, self-contained web application designed for remote language assessment. The frontend was developed using Angular (https://angular.io/), with a NodeJS backend (https://nodejs.org/en/), for our data collection we deployed Lingo on Amazon Web Services (AWS, https://aws.amazon.com/). We acknowledge other researchers may have challenges using AWS to deploy Lingo, so we also provide a pre-compiled version of Lingo that can be run locally. This modular architecture enables straightforward customization of task batteries and scalable deployment.

Upon accessing the platform URL generated once Lingo is deployed, participants undergo automated device compatibility testing to ensure audio recording and playback capabilities. The complete battery requires approximately 25-35 minutes to complete, with timing dependent on individual performance in adaptive tasks (matrix reasoning and listening comprehension). All sessions capture three data streams simultaneously: raw audio recordings, response selections, and precise timing measurements.

### Lingo Task Descriptions

#### Controlled Oral Word Association Test (COWAT)

The COWAT (also called COWA or FAS) is a well established verbal fluency and executive functioning task [49, 50]. In this task, we asked participants to say as many words beginning with a specified letter as they could for 30-seconds. Participants completed 5 trials total, with each trial being a different letter (A, C, F, L, S). Audio recordings for each trial captures all vocalizations that can be used analysis of word count, response timing, and linguistic features.

#### Sentence Repetition

Sentence repetition is a particularly important task in language assessment, it has previously been shown to be a robust predictor of overall language ability and clinically relevant language disorders and is included in clinical language assessments like the CELF-5 [51, 52]. Our version was adapted from clinical language assessments protocols, and augmented with additional high-difficulty sentences to prevent ceiling effects. Participants were presented with audio of a sentence, hearing the prompt sentence only once, and after a 3-second delay they are asked to repeat what they heard and audio of their response is recorded. This was repeated for a total of 12 trials, with each trial increasing in difficulty (prompt sentences can be seen in S4). For example, the first prompt is “Didn’t the girl paint the picture”. The final prompt is: “Having been tardy more days than not, the brothers both knew it was only a matter of time before each would have to be responsible for himself.”

#### Rapid Automatized Naming (RAN)

The RAN task was included for its relevance to reading skills, previous work has shown RAN performance is strongly related to reading fluency and dyslexia [27, 53]. Following established RAN protocols, we presented participants with a grid of numbers and asked them to read the numbers aloud as rapidly and accurately as possible. Participants completed four RAN trials (two 25-item 5×5 grid trials and two 36-item 6×6 grid trials; all prompts shown in Figure S5). Audio recordings allow for measuring reading speed, error rates, types of errors (e.g., skipping a line or repeating numbers), and timing of errors.

#### Sync Voice (rhythm)

Lingo also includes a rhythm keeping task (called sync voice), while less established than our other tasks, rhythm is a key component of language and has shown robust genetic associations [7, 8, 54]. In this task, participants were asked to continue a rhythm given in an audio prompt. Participants would hear “la” at different timing intervals (0.5, 0.75, 1.0, or 1.25 seconds) for 5 seconds, and after a 3 second delay participants were asked to continue the rhythm. Essentially, participants were asked to say “la la la” at a provided rhythm. Audio files can be analyzed to extract timing information.

#### Picture Narration

Picture narration tasks, sometimes called picture descriptions, are a useful tool to assess expressive language and spontaneous speech production skills. Previous work has shown this task has relevance to psychiatric conditions and neurodegenerative disorders [55, 56]. We implemented this task by presenting participants with a picture and asking them to describe what they saw for 30 seconds. Participants completed a total of four trials, with each trial consisting of a different picture stimuli. Audio recordings can be used to extract measures including total word count, grammatical complexity, semantic content, coherence, and intonation.

#### Following Directions

To assess receptive language skills we also include a “Following Directions” task, similar tasks are used in clinical language assessments like the CELF-5 [57, 58]. In this task, participants hear a set of directions once (for example, “select the blue triangle underneath the red square”; example shown in Figure S6) and respond by clicking target shapes in visual arrays. Eight trials of increasing complexity assess comprehension of spatial-relational instructions. Both the response and time to respond are recorded for each trial. Notably, we found this task had the lowest ceiling across all of our assessments. Future research may want to include more challenging prompts to increase individual variability in these scores.

#### Matrix Reasoning

Given the emphasis on language abilities in the other 6 Lingo tasks, we include a matrix reasoning task to measure nonverbal abilities. This task is primarily involves nonverbal pattern recognition. With permission from the UK Biobank, we used their well validated fourteen visual-spatial reasoning problems in Lingo [59]. Each trial presents an incomplete pattern matrix with six response options, participants select the option that completes the pattern (example shown in S7). Similar to the “Following Directions” task, responses and time to respond are recorded.

### Audio Processing and Transcription

All audio recordings from Lingo tasks (Sentence Repetition, RAN, Picture Narration, and COWAT) underwent automated transcription using WhisperX (large-v3 model) [60]. WhisperX is a time-aligned speech recognition system that provides both transcription accuracy and temporal alignment of utterances.

To validate transcription accuracy of WhisperX on our data, we manually transcribed a random sample of 52 COWAT audio files. Manual transcription was performed blind to the WhisperX output by a trained researcher (L.G.C.) who listened to each file an average of 10 times to ensure high confidence in the transcriptions. Comparison of manual and WhisperX transcriptions revealed 95% concordance at the word level, substantially higher than the 85-88% concordance observed with earlier Amazon transcription services that were evaluated but not used in the final analysis. Based on this high concordance rate, we used WhisperX transcriptions for all subsequent analyses.

To limit potential transcription hallucinations (where the model generates words not actually present in the audio), we filtered transcriptions to include only words with spoken word time *>* 0.1 seconds and detected language of English.

Word counts were automatically extracted from the validated transcriptions and used as primary features for the Picture Narration and COWAT tasks.

### Participant Recruitment and Data collection

#### SPARK Research Match Cohort

We recruited 2,746 total adults through the SPARK Research Match platform, of these 2,293 completed all assessments [25, 26]. Participant demographics can be seen in Figure S2. Exclusion criteria included self-reported blindness or deafness due to task requirements involving visual and auditory stimuli.

#### Test-Retest Reliability Subsample

A subset of 111 participants completed the Lingo battery twice, separated by a minimum two-week interval, to establish test-retest reliability. Test-retest reliability was calculated using Pearson correlations and intraclass correlation coefficients (ICC) on factor scores. To test whether performance between the two sessions significantly improved, we conducted paired t-tests for each factor.

#### Clinical Assessment Validation Cohort

Twelve participants from the SPARK sample with available Wechsler Intelligence Scale assessments enabled validation of Lingo-derived factor scores against clinical measures of cognition (Full Scale IQ, Verbal IQ, Nonverbal IQ) [61].

#### Additional SPARK Phenotypes

Self-reported language impairment and dyslexia used in the comparative power analysis was reported after completing Lingo as part of the SPARK Research Match study. Participants were given the option to self-report a variety of language related questions, including questions about learning disorders (like language impairment and dyslexia) and psychiatric conditions (N = 1,283 who completed the questionnaire). Additional SPARK phenotype data (self-reported ASR scores and parent reported VABS-3 Communication Scores for independent adult with autism) came from the SPARK V14 phenotype release (12-17-2024) [25].

### Data Quality Control and Participant Exclusions

#### Participant Inclusion Criteria

Participants were initially enrolled if they: (1) were at least 18 years old, (2) self-reported English as their primary language, and (3) successfully initiated the Lingo assessment through the SPARK web portal.

#### Quality Control Procedures

##### Step 1: Task Completion

A total of 2,746 participants initiated Lingo. Participants who failed to complete the full battery were excluded from all analyses (N = 241 who started Lingo but stopped before completing all assessments). Primary reasons for non-completion included: technical difficulties (e.g., microphone access issues, browser incompatibility) and voluntary withdrawal before task completion.

##### Step 2: Audio Quality Assessment

Audio recordings were assessed for quality issues that would preclude accurate transcription or feature extraction, for example if the audio file was not as long as it should be that would indicate the participant left before completing the trial or a technical issue. Additionally, if a participant completed Lingo but did not speak during the tasks they would also be excluded. A total of N = 212 participants were excluded due to poor audio quality/non-engagement.

##### Step 3: Item-Level Feature Quality Control

We extracted 41 item-level features from Lingo tasks (Table S1). To ensure data quality, we applied the following filters:

(1) Low variance items: We removed items with low phenotypic variance (variance *<* 0.05).
(2) Floor and ceiling effects: Items were removed if they showed obvious ceiling or floor effects, defined as less than 15% of participants answering incorrectly or more than 85% answering correctly, respectively.

This criterion led to the exclusion of 5 Matrix Reasoning items, 5 Following Directions items, and 7 Sentence Repetition items from subsequent factor analysis. These items primarily represented very easy tasks (e.g., the first few sentences in Sentence Repetition) or very difficult items (e.g., the final Matrix Reasoning problems) that provided limited information about individual differences in the typical range for the factor analysis. It is important to note that these items may be useful in other populations to detect extreme language abilities.

##### Step 4: Participant-Level Outlier Detection

After computing factor scores (see Factor Analysis section), we identified extreme outliers at the participant level. Participants were flagged as outliers if any of their factor scores exceeded the median *±* 4 times the median absolute deviation (MAD), a measure of variability less sensitive to extreme values than standard deviation. This procedure identified 14 participants whose factor scores suggested anomalous performance patterns. To understand the nature of these outliers, we manually reviewed their audio recordings. This review revealed that many of these participants were not fully engaged with the tasks, with clear evidence of distraction (e.g., watching television, talking to others) during task completion. These 14 participants were excluded from all downstream analyses.

##### Final Sample

After applying all quality control procedures, the final analytical sample included N = 2,279 adults for the factor analysis and all subsequent analyses. The flow of the participants can be seen in Figure S1.

### Lingo Feature Extraction

#### Performance Metrics for Verbal Language Tasks

Lingo’s design and emphasis on audio recording allows for a wide variety of data to be extracted to measure participant performance. For the purposes of this manuscript, we focused on a relatively simple set of features in order to establish validity and reliability of Lingo. For the COWAT and picture narration tasks we counted the number of total number of words said, and delay time (time to say first word). For the sentence repetition task we measured performance with bigram accuracy, the proportion of correct consecutive word pairs in responses relative to target sentences. This continuous scoring approach provides greater sensitivity than traditional 0-3 point scales often used in clinical assessments, particularly for detecting subtle individual differences [58]. For the sync voice (rhythm) task we used the timing of the la’s said by participants to calculate consistency (measured by fitting a regression model for each participant predicting expected timing from actual timing and extracting the *r* ^2^ value - with higher values indicating high consistency) and accuracy (difference in actual rhythm timing and prompt timing). For the RAN task we derived a “reading speed” score, which was simply the time to complete reading aloud the number grid. Future work should incorporate acoustic and semantic features in these tasks.

#### Performance Metrics for Item Selection Tasks

Both the following directions and matrix reasoning tasks do not record any audio, participants are presented with multiple choice questions that ask them to select the correct response. These tasks record both the time to respond and the response.

### Phenotype Imputation

Early in participant recruitment there was an issue with data loss, not all responses to the matrix reasoning and following directions items were recorded correctly. For example, a participant may have completed all of Lingo but would be missing responses for prompts 4, 6, and 11 from the matrix reasoning. This issue specifically impacted the matrix reasoning and following directions tasks, there did not appear to be issues with recording data from any of the verbal language tasks. We found this was likely due to an issue with our AWS deployment (using too small of a server for the number of participants we recruited). After noticing the issue and switching to a larger AWS server we did not have any problems with data loss. Given that all participants had completed the rest of Lingo and had at least some data for each task recorded (missing *<* 1/3 of Lingo items), we opted to impute the missing data. We utilized a tree based approach with predictive mean matching to impute the missing data, with the missRanger R package (N = 1,000 trees, and predictive mean matching k = 5) [62].

### Factor Analysis

#### Exploratory Factor Analysis

We conducted an exploratory factor analysis on 41 quality-controlled features from 2,293 participants after residualizing item level data for age and sex effects. Data factorability was confirmed via Bartlett’s sphericity test (p *<* 2×10*^−^*^16^) and Kaiser-Meyer-Olkin criterion (KMO = 0.88). The four-factor solution was identified through data-driven parallel analysis using the R package psych, which compares observed eigenvalues to eigenvalues of random data with the same dimensionality [63]. We used the standard options in the psych fa function, which uses the regression method, minres factoring method, and oblimin rotation. While the TLI (0.80) was below conventional thresholds for an acceptable fit, the RMSEA (0.051) and SRMR (0.04) values indicated a good fit. The four factor solution explained 33.1% of variance in the data. Importantly, the resulting factors demonstrated strong psychometric properties: with significant test-retest reliability, correlation with clinical Wechsler IQ assessments, and correlations with cognitive polygenic scores. This pattern of external validity supports the utility and interpretability of this factor solution. If participants completed 2 separate Lingo assessments (for test-retest reliability), we only included their first assessment in the factor analysis and predicted their factor scores for their second assessment using the factor solution.

### Polygenic Score Analysis

#### Genotype Quality Control and Processing

Genetic analyses utilized SPARK genotyping data, including array and sequencing genotypes from the iWES1 and iWES3 releases and whole-genome sequencing releases 2-4. For each dataset, we applied standard quality control procedures, filtering variants with missingness *>* 5% and samples with missingness *>* 5%. Sample-level heterozygosity outliers were identified using F-statistics and excluded if they fell beyond 3 standard deviations from the mean. Strand alignment was verified against the TopMed reference panel to identify and correct strand flips.

#### Genotype Imputation

Genotypes from the iWES1 and iWES3 genotypes were imputed to the TopMed reference panel. We retained variants with imputation quality scores *r* ^2^ *>* 0.8 and minor allele frequency (MAF) *>* 0.1%. Following imputation of the iWES1 and iWES3 genotypes, we performed liftover from hg38 to hg19 coordinates, normalized variant representations to the reference genome, and merged data across sequencing batches. The final dataset comprised 1,227,051 HapMap3+ variants for polygenic score calculation.

#### Population Stratification

We performed principal component analysis using bigsnpr to characterize population structure [64]. We computed genetic principal components on unrelated samples (calculated using KING, relatedness cutoff = 2^-3.5^) using the bed autoSVD function that removes rare varaints and prunes variants [65]. Genetic outliers were identified and removed using protocols recommended by the package author, and remaining samples were projected onto principal components identified from the unrelated individuals.

#### Polygenic Score Construction

We constructed polygenic scores (PGS) for 15 traits using LDpred2-infinitesimal, leveraging linkage disequilibrium (LD) estimates from the provided UK Biobank European ancestry reference panel. Target traits spanned multiple domains: psychiatric conditions (ADHD [66], autism spectrum disorder [67], bipolar disorder [68], major depression [69], schizophrenia [70], PTSD [71], anorexia nervosa [72], alcohol use disorder [73]), cognitive abilities (cognitive performance [74], educational attainment [9], executive function [75]), neurological conditions (epilepsy [76], rare neurodevelopmental conditions [77]), and behavioral traits (risk-taking behavior [78], addiction liability [79]). To control for population stratification, we residualized each PGS for the first 20 genetic principal components prior to analysis. All PGS were then z-score standardized (mean = 0, SD = 1) to enable comparison of effect sizes across traits. Finally, we filtered to samples that were in the Lingo Research Match (N = 2,098).

#### Association Testing Between Polygenic Scores and Lingo Factors

We tested associations between each PGS and each Lingo factor using Pearson correlations (60 total tests: 15 PGS *×* 4 factors). To account for multiple testing, we applied False Discovery Rate (FDR) correction using the Benjamini-Hochberg procedure across all 60 tests. Associations with FDR-adjusted p *<* 0.05 were considered statistically significant. Full summary statistics can be found in Table S8.

#### Comparative Power Analysis Framework

To quantify the statistical efficiency of Lingo-derived phenotypes relative to traditional language assessments, we conducted a comparative power analysis across four measurement approaches: (1) Lingo *g* factor (F3) scores, (2) VABS-3 Communication Standard Scores, (3) self-reported dyslexia diagnosis, and (4) self-reported language impairment diagnosis. The VABS-3 (Vineland Adaptive Behavior Scales, 3rd Edition) Communication domain is a clinically validated questionnaire commonly used to assess language and communication skills in individuals with developmental and intellectual disabilities. We used the cognitive performance PGS as a benchmark effect, as it shows robust associations with language-related phenotypes in prior literature. For each phenotype and sample size combination, we performed bootstrap resampling with 1,000 iterations. In each iteration, we randomly sampled participants (with replacement), regressed the phenotype on cognitive performance PGS while controlling for age, sex, and the first five genetic principal components, and recorded whether the PGS association achieved nominal significance (p *<*0.05). Statistical power was defined as the proportion of 1,000 bootstrap iterations yielding significant associations.

We evaluated sample sizes ranging from 100 to 1,500 in increments of 25 participants. For self-reported diagnoses, we extrapolated beyond 1,500 participants to estimate the sample size required for 80% power, as observed power plateaued near 60% even at N = 1,500. This extrapolation assumed the empirical relationship between sample size and power would continue at the observed rate. To ensure fair comparison, several steps were taken: (1) VABS-3 analyses were restricted to independent adults with autism in SPARK, excluding children and dependent adults, as this subsample was most comparable to the Lingo sample; (2) all participants in the self-report diagnosis analyses had also completed Lingo, ensuring identical sample composition; (3) identical covariate sets and statistical approaches were applied across all phenotypes. We note that including all available VABS-3 participants (including children and dependent adults) substantially reduced power to detect cognitive performance PGS associations, likely reflecting greater phenotypic heterogeneity and measurement error in the excluded subgroups.

A summary table of the comparative power analysis results can be found in Table S11.

### Rare Variant Association Analysis

#### Variant Annotation and Filtering

Pre-processed whole-exome sequencing data came from SPARK’s iWES v3 dataset, with variants called using GATK and filtered with strict quality control [25, 80]. Variants were annotated with the Ensembl Variant Effect Predictor tool (VEP v109 using the hg38 reference) [81] and FAVOR [82] to add allele frequencies, predicted variant effects, mapping variants to genes, and deleteriousness predictions (CADD scores) [83]. We identified rare variants based on both reference population allele frequencies provided by VEP and SPARK allele frequencies (max reference population MAF *<* 1% and SPARK MAF *<* 1%).

#### pLI Gene Analysis

For the pLI analysis, we used all genes with a pLI *>* 0.9 based on gnomAD v4.1 and counted up the number of genes where each sample carried a rare predicted loss-of-function variant or missense variant [84, 85]. pLI gene burden was calculated as the number of rare predicted loss-of-function variants and missense variants across all protein coding genes with a high pLI score, we also computed genome wide burden to test whether the effects are specific to pLI genes or general burden. Analyses controlled for population stratification using the first 5 genetic principal components and ancestral superpopulation, genome-wide rare variant burden (sum of CADD scores across all protein coding genes) to account for individual mutational load differences, polygenic score backgrounds (cognitive performance PGS) to separate common from rare variant effects, and demographic covariates including age and sex.

#### Gene-Level Association Testing

Given the underpowered sample size for genetic discovery, we opted to use gene-level tests instead of variant level tests. To improve power, our approach utilized a quantitative approach to measuring gene burden, rather than a binary indicator of variant class that is more commonly used. We used the maximum CADD score that that each individual carries in a gene for association testing [83]. For example, if someone carries two rare variants in a gene and one had a CADD score of 5 and another of 25, we would use the 25 as their quantitative measure of burden in that gene. If someone did not carry a rare variant in a gene, they would get a burden score of 0. We limited our analysis to genes with at least 20 individuals carrying a rare variant in that gene and genes where someone in our sample carried a variant with a CADD phred score *≥* 20 (total genes tested = 13,760). Similar to the pLI association testing, all gene-level analyses controlled for population stratification using the first 5 genetic principal components, ancestral superpopulation, genome-wide rare variant burden (sum of CADD scores across all protein coding genes) to account for individual mutational load differences, polygenic score backgrounds (cognitive performance, educational attainment, executive functioning, autism, ADHD, and rare neurodevelopmental condition PGSs) to separate common from rare variant effects, and demographic covariates including age and sex. Given the large number of statistical tests, we corrected p-values using the Bonferroni method to reduce type 1 errors, which gave a stringent p *<* 3.6 *×* 10*^−^*^6^ cutoff for statistical significance (0.05 / 13,760 genes tested). Genome-wide association summary statistics are provided in Table S10.

#### Pathway Enrichment Analysis

Following the gene-level association testing, we examined enrichment of biological pathways and gene sets using Gene Ontology and GWAS Catalog. We grouped all nominally significant genes associated with any of the Lingo factors (all genes with an unadjusted p *<* 0.05) and tested for enrichment using the enrichR R package [86]. Similar to the gene-level association testing, we used Bonferroni corrected p-values to determine statistical significance and reduce type 1 errors.

#### Candidate Gene Validation with MAGMA

In order to provide independent validation of our candidate genes (*NGB* and *GLS*) we examined gene-level association results using MAGMA [87] on large scale common variant GWAS results for educational attainment [9] and dyslexia [5]. We used the most recent educational attainment GWAS summary statistic, and ran MAGMA using standard parameters (mapping SNPs to genes based on location, using the recommended 35Kb or less upstream and 10Kb or less downstream cutoffs). The dyslexia GWAS summary statistics are not publicly available, so we utilized the MAGMA results from the publication’s supplementary tables.

### Mental Health Association Analysis (ASR)

Linear regression models assessed relationships between Lingo factors and ASR subscales, controlling for Total Problems T-scores, age, and sex to identify specific symptom domain associations beyond general psychiatric distress. This approach enabled identification of factor-specific psychiatric associations while accounting for overall mental health burden. To account for multiple testing, we applied False Discovery Rate (FDR) correction using the Benjamini-Hochberg procedure across all tests. Associations with FDR-adjusted p *<* 0.05 were considered statistically significant. Full summary statistics can be found in Table S7.

### Software and Reproducibility

All analyses were conducted in R version 4.3.0 [88]. The Lingo platform code is publicly available at: https://research-git.uiowa.edu/michaelson-lab-public/lingo.

#### Human Subjects Approval and IRB

Institutional Review Board approval was obtained from the University of Iowa for the Research Match study (201705739), SPARK is approved through the WCG Institutional Review Board (formerly known as Western IRB; 201703201). All participants provided informed consent for genetic and phenotypic data collected and used in this study.

## Supporting information

All supplementary tables

## Acknowledgments

We thank Ben Tysseling for his contributions to the Lingo codebase and platform implementation. We are grateful to all of the participants and families in SPARK, the SPARK research match team and Tempus, the SPARK clinical sites, and SPARK staff. We appreciate obtaining access to genetic and phenotypic data for SPARK data on SFARI Base.

We are especially grateful to Dr. Bob McMurray, whose thoughtful feedback on an earlier version of this manuscript greatly improved our approach to psychometrics and data analysis.

## Data availability

The SPARK genetic and phenotypic data can be obtained at SFARI Base: https://base.sfari.org The Lingo data collected as part of the SPARK Research Match is available to qualified, approved researchers through SFARI Base upon request.

## Author contributions

Lingo was originally conceived by JJM with input from JBT and the initial version developed by MS. Further conceptual and software development was contributed by TK, KM, and LGC. The study was designed by LGC, TK, and JJM. The SPARK Research Match study was carried out by TK and LGC with guidance by JJM. The data was processed by LGC, TK, and ME. The analyses were performed by LGC, TK, ME, YH, EK, GS, TJ,and JJM. The manuscript was drafted by LGC and JJM and was approved by all authors.

## Supplementary information

### Supplementary tables and figures

**Figure S1.**
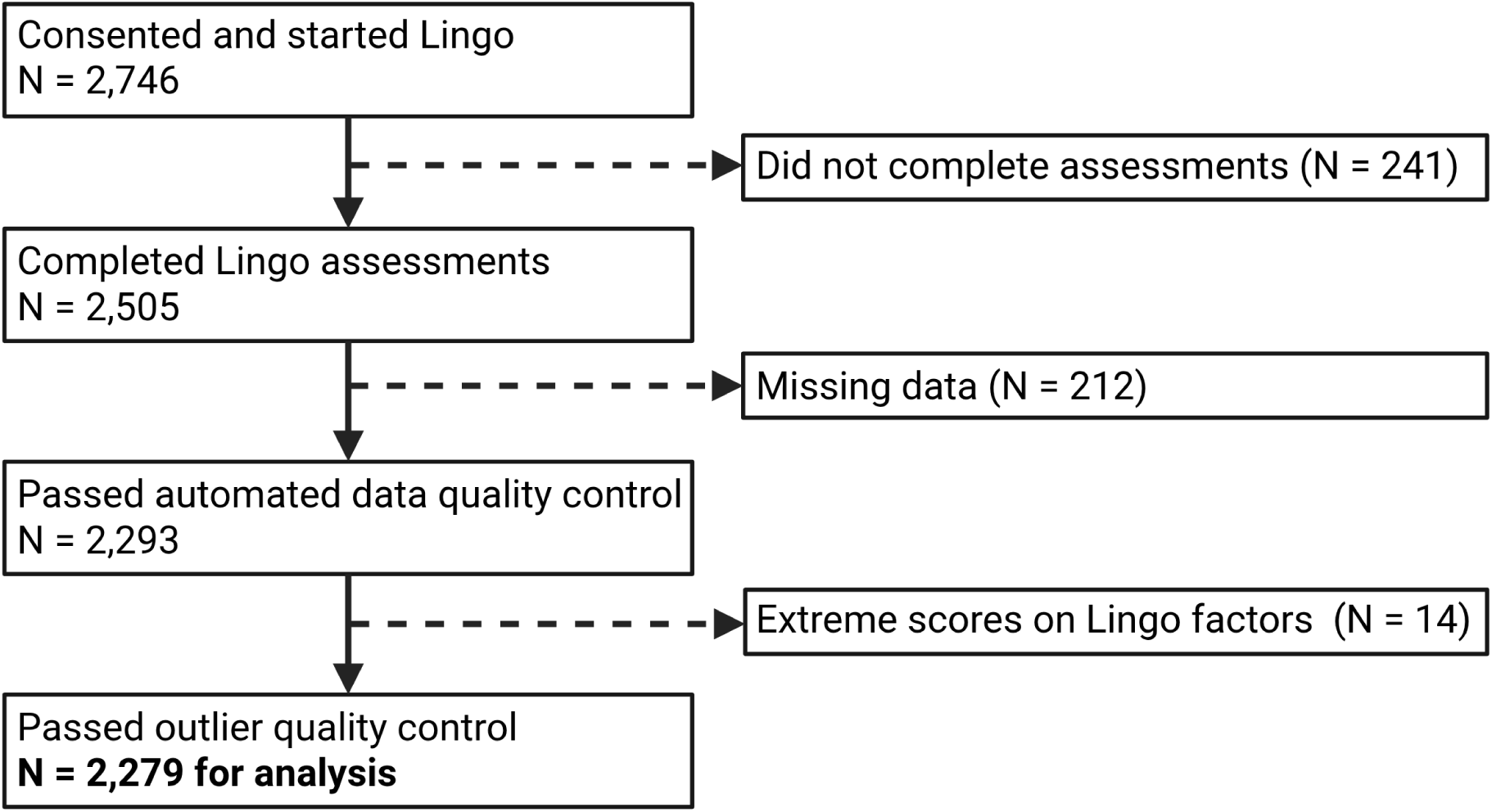
Participant enrollment and quality control flowchart. Flow diagram showing participant progression through Lingo assessment and quality control procedures. Boxes on the left show the main flow of participants from initiation through final analytical sample. Boxes on the right indicate exclusions at each stage. Diagram was made in BioRender.

**Figure S2.**
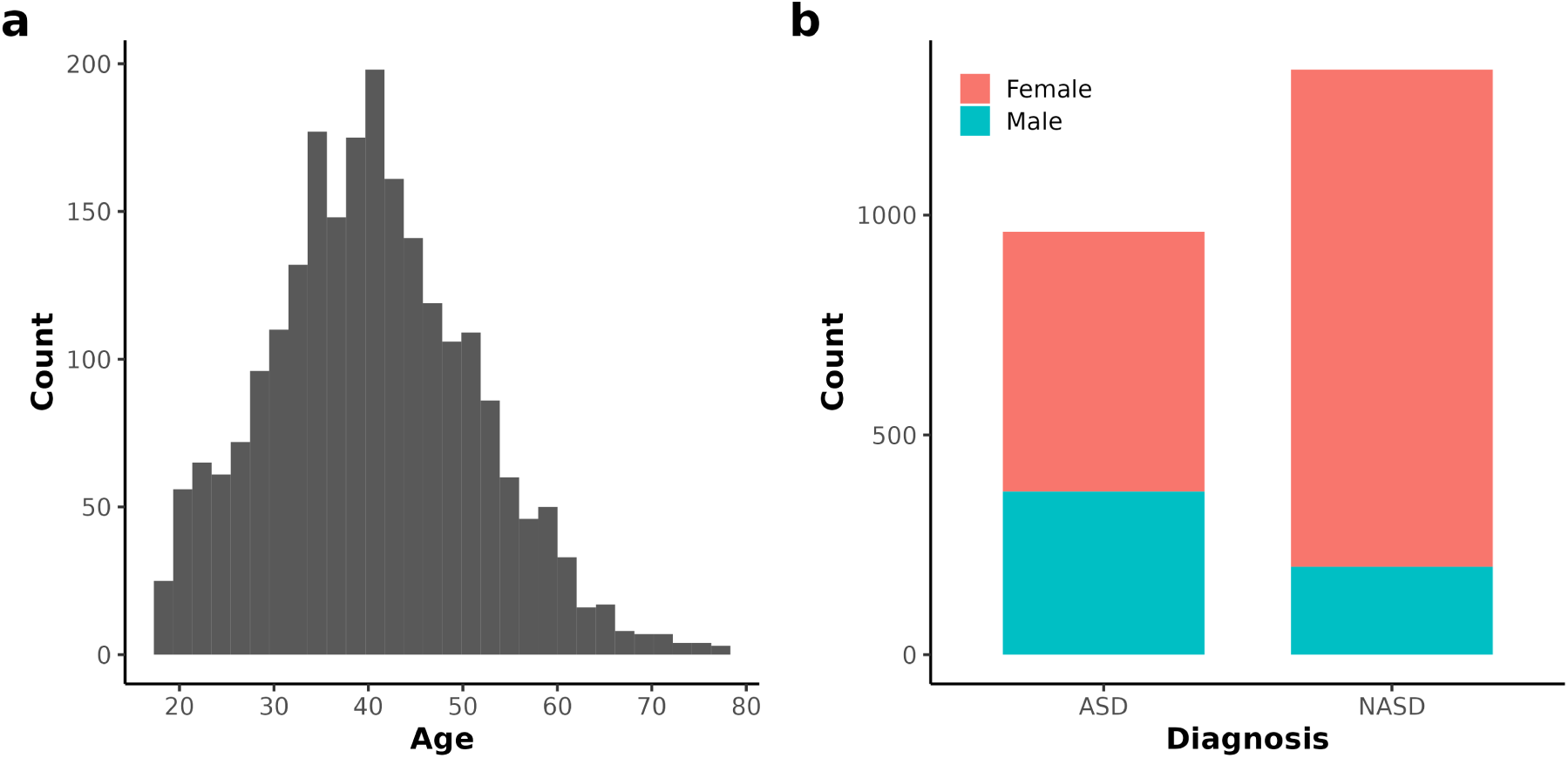
Demographics of the SPARK Lingo participants. **a** Distribution of age across individuals who completed Lingo in the SPARK Research Match study (N = 2,293). **b** Count of individuals with and without autism in the sample, colored by sex.

**Figure S3.**
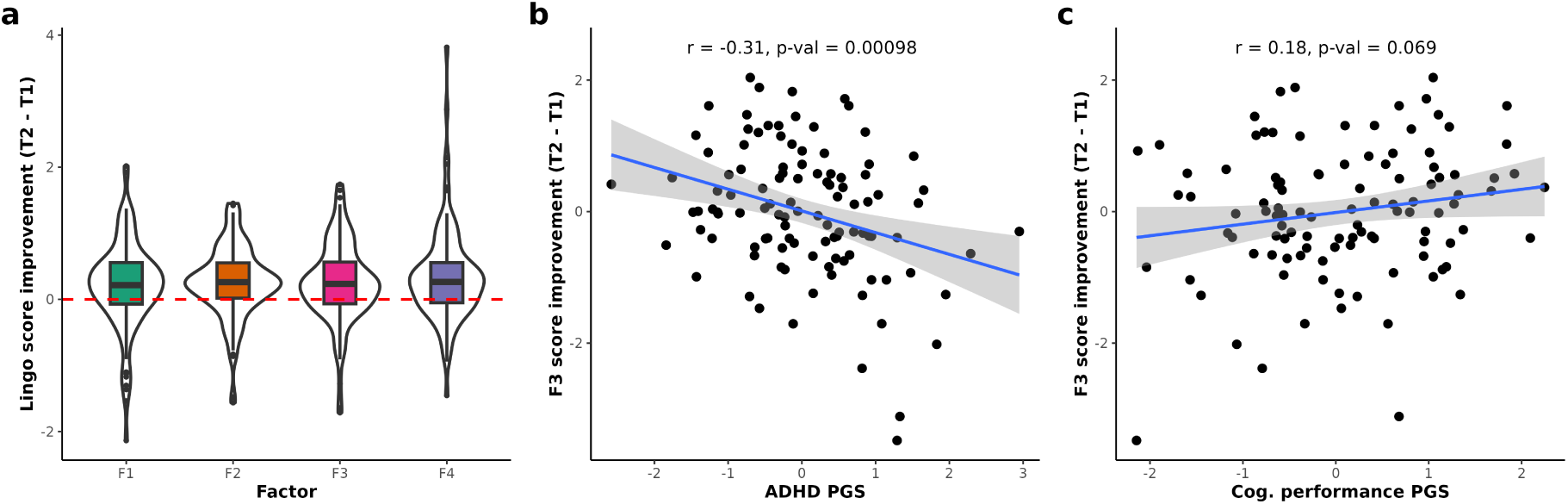
Lingo learning effect and genetic associations. (**a**) Distribution of improvement scores (T2 - T1) across the four Lingo factors, showing consistent performance gains across all dimensions. (**b**) Scatterplot showing a negative correlation between *g* (F3) factor score improvement and ADHD polygenic scores. (**c**) Scatterplot showing a trending positive correlation between *g* (F3) factor score improvement and cognitive performance polygenic scores.

**Figure S4.**
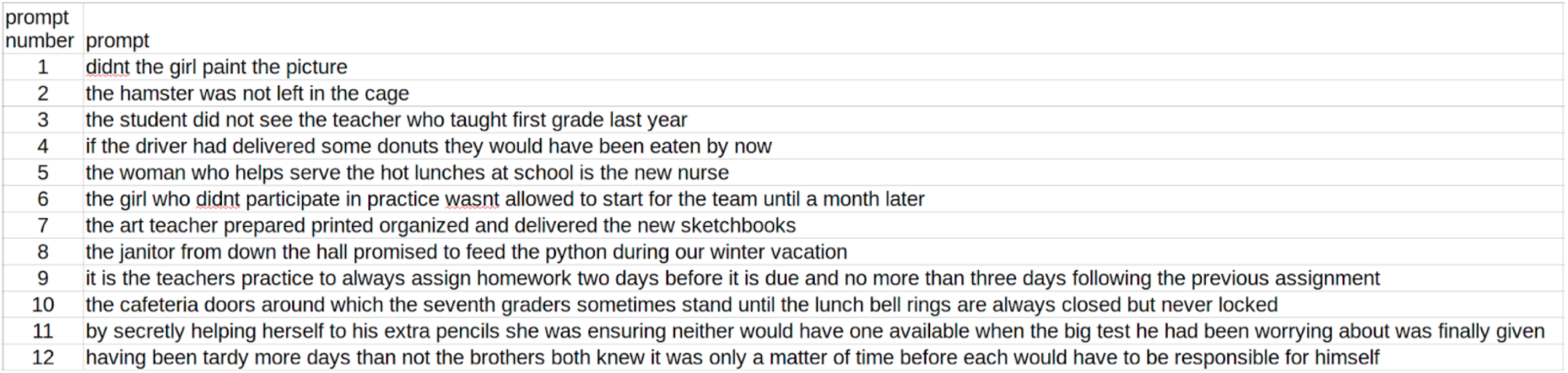
**Prompt sentences used in the sentence repetition task (with punctuation and capitalization removed).**

**Figure S5.**
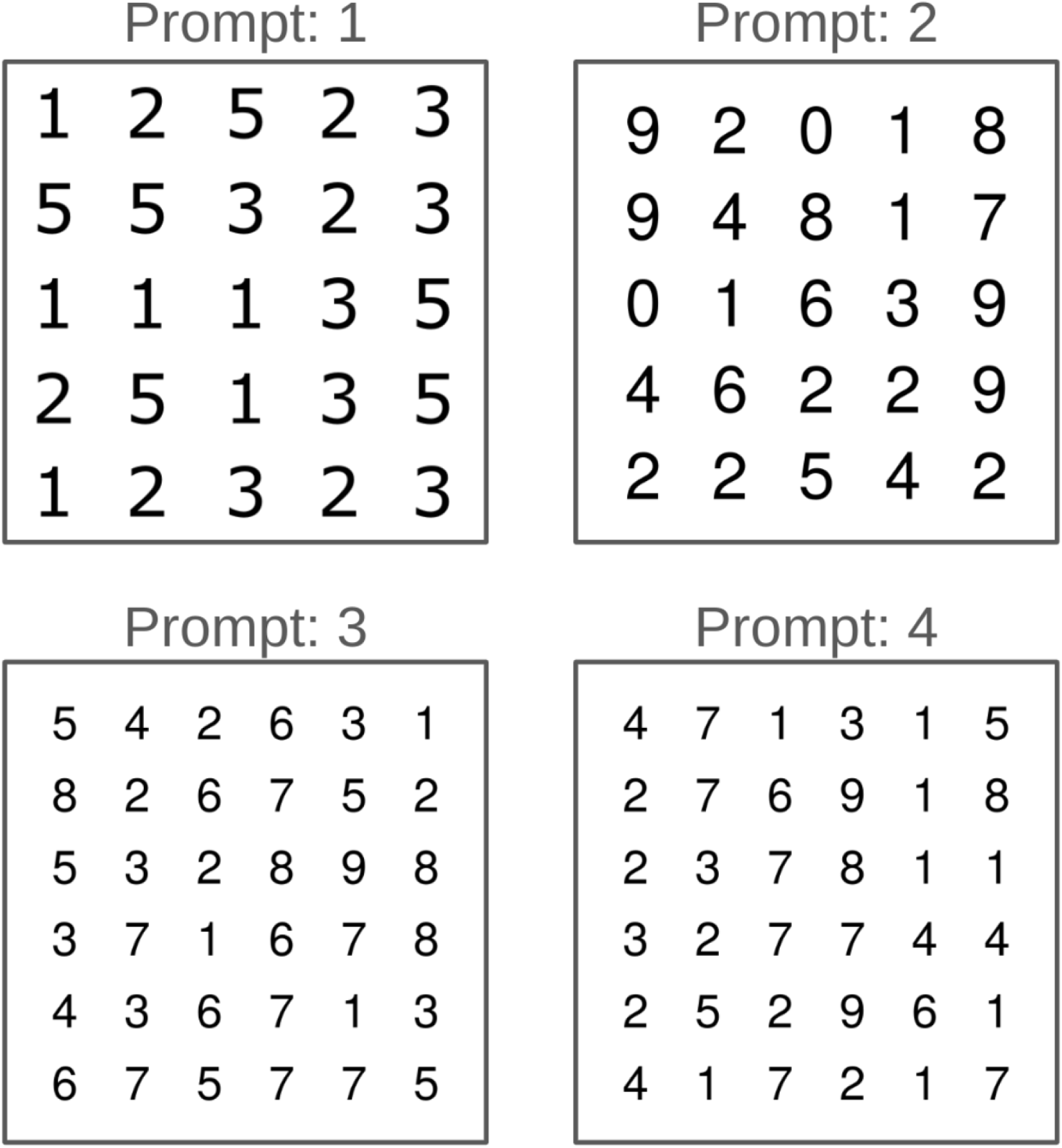
**Prompt number grids used in the RAN reading task.**

**Figure S6.**
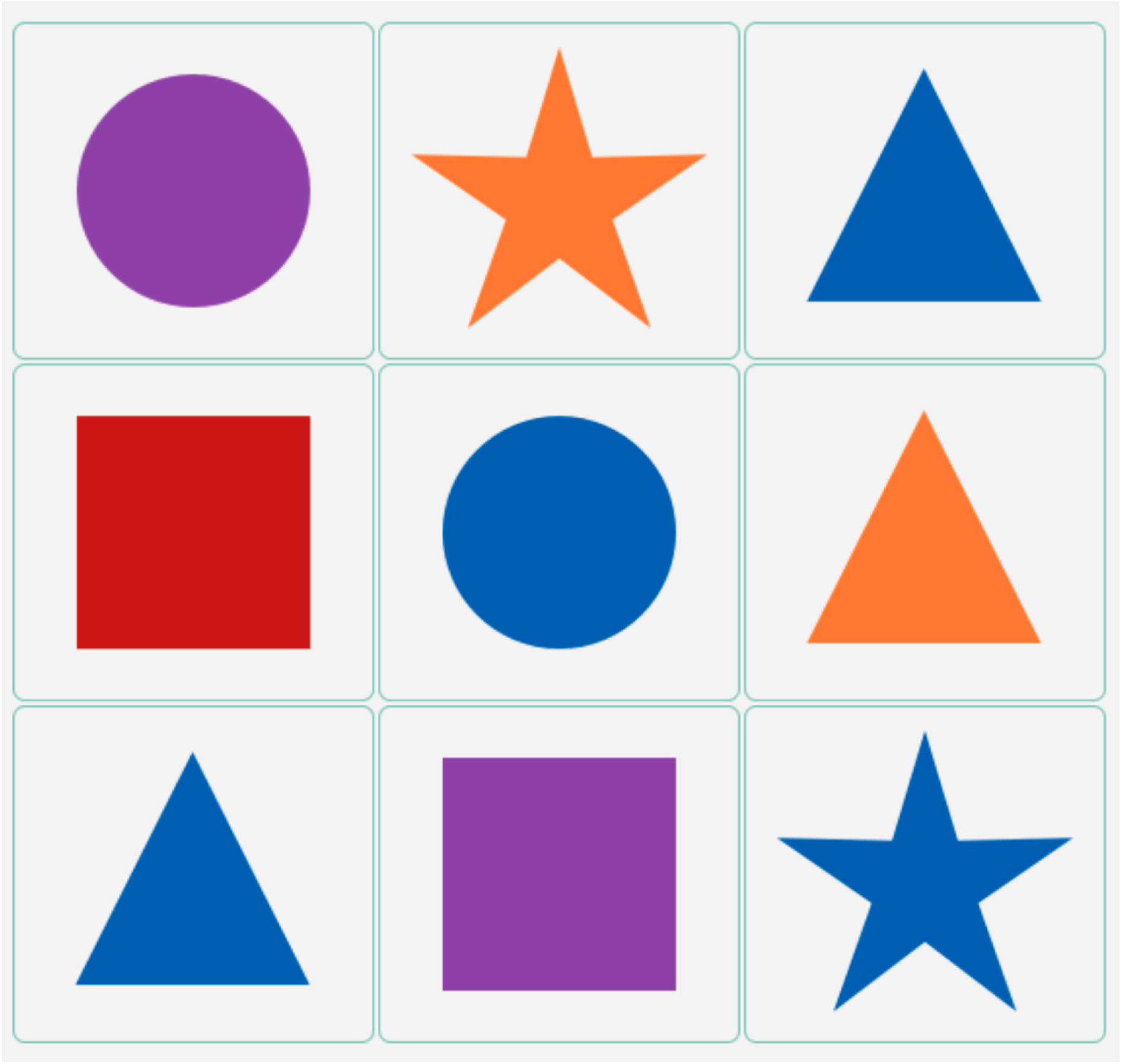
Prompt 1 image for the following directions. Participants were asked to click on the shape that matched the verbal description they had heard.

**Figure S7.**
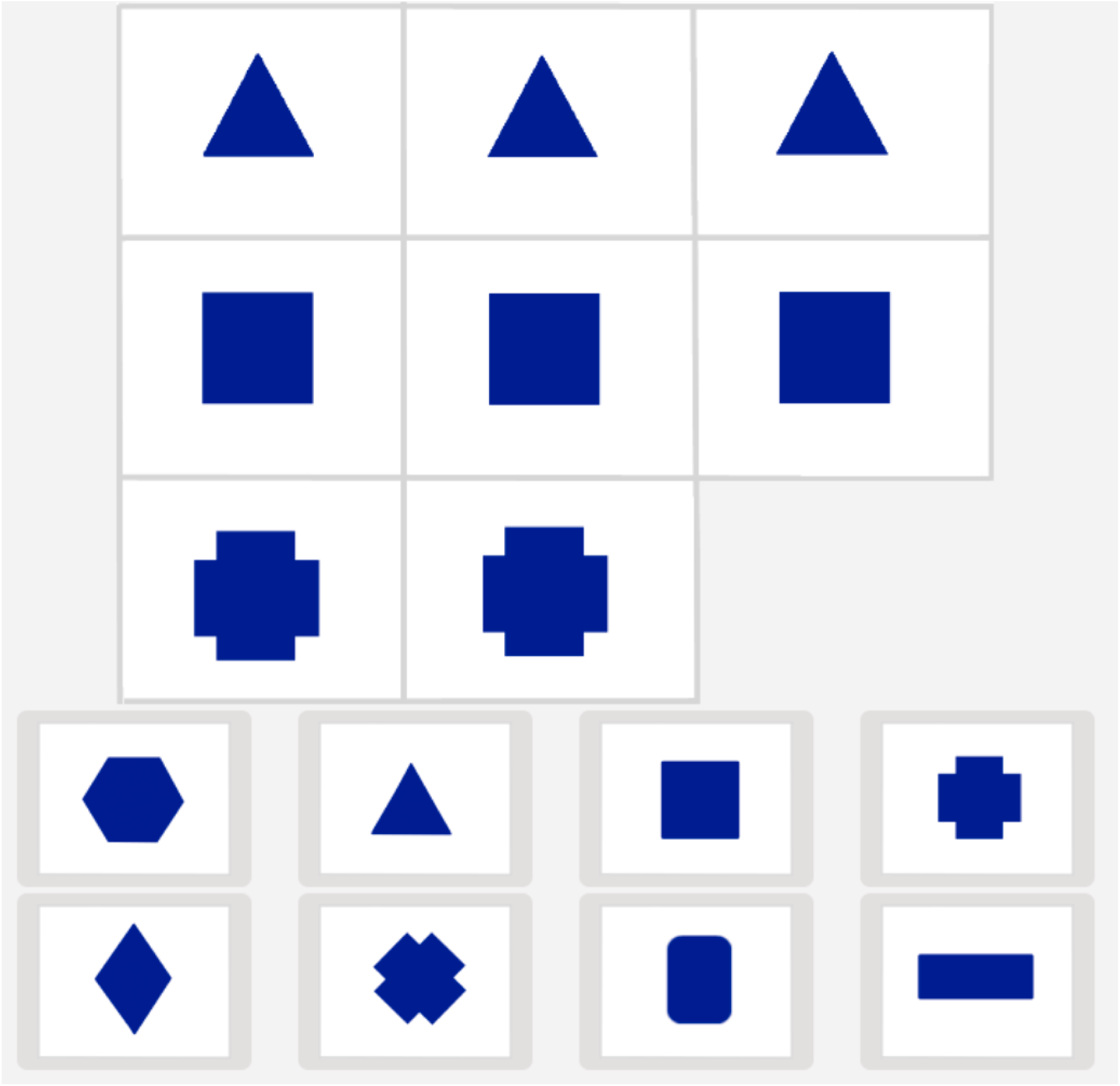
Prompt 1 image for the matrix reasoning task. Participants were asked to click on the pattern that best fit the matrix shown above.

